# A penalized distributed-lag non-linear model for modeling the joint delayed effect of two predictors: impact of minimum and maximum temperature on mortality

**DOI:** 10.1101/2024.11.29.24318041

**Authors:** Sara Rutten, Elisa Duarte, Thomas Neyens, Dirk Lauwaet, Christel Faes

**Affiliations:** I-BioStat, Data Science Institute, Hasselt University, Hasselt, Belgium; L-BioStat, Department of Public Health and Primary Care, KU Leuven, Leuven, Belgium; VITO – Flemish Institute for Technological Research, Mol, Belgium

**Keywords:** Distributed lag, Interaction effects, Nonlinear model, Spatial risk-analysis, Temperature, Time series

## Abstract

Distributed lag non-linear models (DLNMs) offer a flexible approach towards modelling time-delayed exposures. They are popular to study the effect of environmental exposure on health outcomes, such as the effect of temperature on mortality. Conventional distributed lag non-linear models typically focus on a single exposure variable, potentially overlooking complex interactions between multiple predictors. In this paper, we propose a distributed lag non-linear model that captures the joint delayed impact of two exposure variables by incorporating their interaction through a tensor basis constructed from univariate P-splines. This model is compared to a model assuming an additive effect of delayed exposures. Our model is used to examine the joint impacts of maximum and minimum temperatures on all-cause mortality in Flanders during summer. The results show that our model provides a flexible strategy towards the analysis of two predictors with interacting time-delayed effects on an outcome of interest. The importance of both maximum and minimum temperatures in explaining variability in mortality is illustrated, and we show that the interaction effect varies across age and gender groups. A spatial risk analysis at the municipality level reveals that mortality is attributed differently to temperature exposure across different areas, due to temperature variations as well as spatial trends in age and gender.

## 1. Introduction

The family of distributed lag non-linear models (DLNMs), proposed by Gasparrini et al. (2010), provides an elegant and flexible approach towards modelling an exposure-response relationship when the exposure effects are lagged. Therefore, the DLNMs have been used extensively in situations where the effect of an exposure persists beyond its initial occurrence. The response *y*_*t*_ is modelled based on past occurrences *x*_*t−l*_ of a predictor variable *x*, where *l* is called the lag. Potential non-linearity in both predictor as well as lag dimension can be addressed by the use of splines. Originally, DLNMs required user selection of the spline parameters, such as the number of knots and placement of these knots. However, Gasparrini et al. (2017) proposed an extension of the DLNM, based on penalized splines. A large number of knots is included in each dimension and a penalty term is added to ensure smoothing and prevent overfitting.

Given the potential time delay in the impact of temperature on health outcomes, the association between extreme heat and mortality risk is an example for which the DLNM is particularly well-suited. Examples can be found in Gasparrini et al. (2015), Bao et al. (2016), Guo et al. (2022), Demoury et al. (2022) and Scovronick et al. (2018), among others. Although temperatures can substantially fluctuate throughout the day, studies often rely on a single summarizing measure of daily temperature, typically either the maximum, minimum or mean temperature.

While most studies are based on maximum temperature or mean temperature, Wei et al. (2021), Murage et al. (2017) and Royé (2017) show that minimum temperature can have an important effect as well, not entirely explained by adjusting for maximum or mean temperature.

Not only the mean, maximum, and minimum temperatures, but also their temporal persistence has been linked to adverse health effects (Anderson and Bell, 2011): heat waves are typically defined based on a minimum number of days with daily maximum temperatures that exceed a certain threshold. In Flanders, the northern part of Belgium, a heat wave is defined as a period with at least 25 degrees Celsius maximum temperature for five consecutive days, with at least three of those days reaching 30 degrees Celsius (KMI). Besides, also Braga et al. (2001) argued that exposure to extreme temperatures can impact health for a period that extends several days beyond the initial exposure. However, as previous studies indicate that maximum as well as minimum temperatures can be associated to adverse health effects, it is important to investigate their potential combined effect in a setting that allows for time-persistent effects. This can be done using a DLNM approach that allows for the joint modelling of two delayed covariates. The traditional DLNM assumes additive affects, but interactions of the effects of maximum and minimum temperatures are plausible, since the combination of hot days and nights can limit the capacity of the body to recover (Besancenot, 2002). To the best of our knowledge, the interaction between these two temperature measures on mortality has not yet been studied. We propose an extension of the traditional DLNM that allows for interacting lagged effects of two predictors. We compare our model with traditional DLNM parametrizations that differ in the way they model the lagged effect of temperature on mortality: by including only maximum temperature, only minimum temperature or the additive effect of both maximum and minimum temperatures.

Section 2 provides an overview of the dataset that is provided by the Department of Care of the Flemish Government. In Section 3, the traditional penalized DLNM model is summarized and an extended DLNM model, allowing to describe the joint delayed effect of two predictors is proposed. The performance of the proposed model is investigated in Section 4 using a simulation study. Section 5 summarizes the results on the impact of minimum and maximum temperature on mortality in Flanders.

## 2. Mortality data in Flanders

The dataset contains daily mortality counts from the year 2000 until 2019 for each of the 300 Flemish municipalities. In addition, it includes population size in different age and gender groups, as well as daily maximum and minimum temperatures. As data is incomplete for Herstappe, the smallest municipality in Flanders, this municipality is excluded from the analysis. The daily minimum and maximum temperature data at the municipality level were calculated from model simulations with the urban climate model UrbClim (De Ridder et al., 2015), in the framework of a project commissioned by the Flemish Environmental Agency. The UrbClim model was applied to simulate hourly temperatures for all warm period (April - September) of the years 2000-2019 with a horizontal resolution of 100m. These simulations were driven by ERA5 reanalysis data and used a detailed local land use map (Poelmans et al., 2022) as input to downscale the ERA5 air temperatures. The model results have been validated against 12 measurement stations across Flanders, yielding excellent error statistics. This study specifically focuses on the summer period, defined as the period between May 15 and September 30. In total, there are 338,726 deaths from any cause that occurred in Flanders within the summer periods from 2000 until 2019. Temperature differences between the 299 included municipalities in Flanders exist (see Figure S1 in the Supplementary Materials). A clear gradient in terms of maximum temperatures is clearly present, with higher maximum temperatures in the eastern part of Flanders and lower maximum temperatures in the western part of Flanders, near the coast. For minimum temperatures, mainly the cities in Flanders have higher temperature values. This aligns with observations that urban areas undergo less cooling at night (Oke, 1982).

The mortality rate in winter, i.e., from November until March, was calculated for each municipality, as it could potentially impact the anticipated summer mortality, i.e. from May until September. Our analysis focuses on two age groups: individuals aged over 85 years and those aged 65 to 84 years. As temperature effects are known to differ between age and gender groups, a stratified analysis is conducted, fitting distinct models for each age and gender subgroup.

## 3. Methodology

### 3.1 The traditional (penalized) DLNM framework

The distributed lag non-linear model of Gasparrini et al. (2010) deals with a non-linear exposure-response relationship as well as non-linear lag relationship. The general model for a single time series of responses *Y*_*t*_ with *t* = 1, …, *n* is given by:

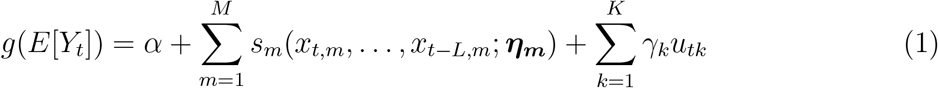

where *Y*_*t*_ follows a distribution that belongs to the exponential family and *g* is the corresponding link function. The *M* exposure variables *x*_*m*_ are related to the linear predictor by using a smoothing function *s*_*m*_ and which allows for a non-linear relationship as well as time-delayed effect. In this, *x*_*t−l,m*_ denotes the variable *x*_*m*_ at time lag *l*, with *L* the maximal lag. The *K* other covariates *u*_*k*_ are linearly related to *g*(*E*[*Y*_*t*_]) with regression coefficients *γ*_*k*_.

Following Gasparrini et al. (2010), defining the smoothing functions *s*_*m*_(*x*_*t,m*_, …, *x*_*t−L,m*_; ***η***_***m***_) consists of two main steps. First of all, a suitable basis for the variable dimension is chosen to allow for a non-linear association between the exposure and the outcome. Let 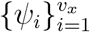 denote the basis function which can be calculated in each value of the exposure variable *x*_*t−l,m*_. Second, a suitable basis for the lag dimension is chosen as well, to account for the temporal correlation between these lagged occurrences. Let 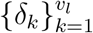 denote the basis function which can be calculated in the lag vector (0, …, *l*, …, *L*)^*T*^. Using this notation, Gasparrini et al. (2010) proposed a distributed lag non-linear smooth term as:

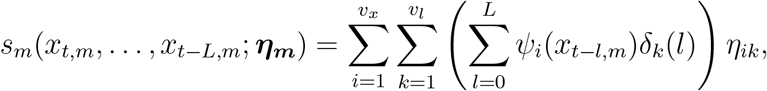

which can be written in matrix notation as:

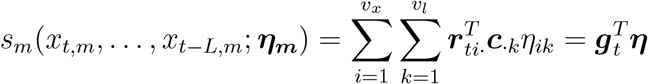

where ***r***_*ti*·_ = (*r*_*ti*0_, …, *r*_*tiL*_)^*T*^, *r*_*til*_ = *ψ*_*i*_(*x*_*t−l,m*_), ***c***_·*k*_ = (*c*_0*k*_, …, *c*_*Lk*_)^*T*^, *c*_*lk*_ = *δ*_*k*_(*l*), ***g***_*t*_ is obtained by applying the *v*_*x*_ · *v*_*l*_ basis functions to *x*_*t−l,m*_ and ***η*** is a vector containing all the *v*_*x*_ · *v*_*l*_ parameters of interest. Note that by including an intercept in the variable dimension, the model in (1) is overidentified. To address this issue, terms associated with the intercept of the variable space are excluded. For consistency in notation, denote by ***η*** the remaining coefficients only.

To mitigate the impact of user-defined specifications on the spline form, P-splines (Eilers and Marx, 1996), a penalized form of B-splines, are used, as demonstrated by Gasparrini et al. (2017). Hence, instead of maximizing the likelihood function *l*(***η, γ***, *α*), the following function will be maximized:

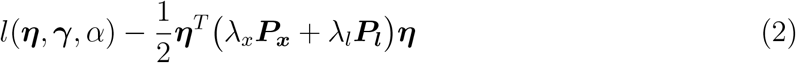

with smoothing parameters ***λ*** = (*λ*_*x*_, *λ*_*l*_)^*T*^. To define ***P***_***x***_ and ***P***_***l***_, we make use of penalty matrices 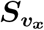 and 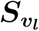, constructed as ***D***_***d***_^*T*^ ***D***_***d***_ with ***D***_***d***_ a difference matrix of order *d*. The number of rows of this difference matrix ***D***_***d***_ is equal to *v*_*x*_ *− d* and *v*_*l*_ *− d* for 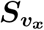 and 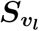 respectively while the number of columns equals *v*_*x*_ and *v*_*l*_ respectively. Examples of such difference matrices are Gasparrini et al. (2017):

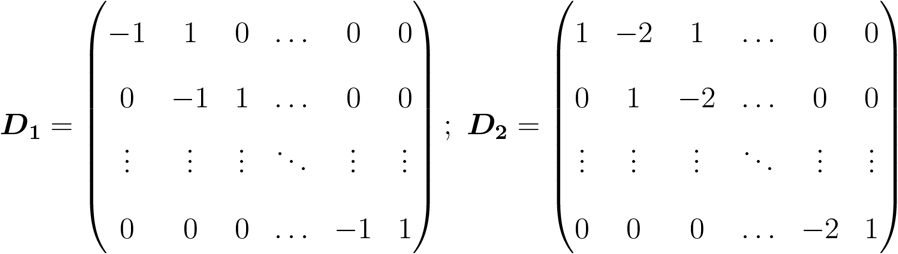

Let 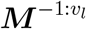 denote the removal of the first *v*_*l*_ rows and columns of a certain matrix ***M***. Accounting for the exclusion of the intercept in the variable dimension, the matrix ***P***_***x***_ and ***P***_***l***_ are then defined as:

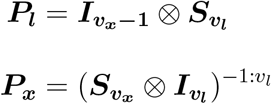

with ***I*** a diagonal matrix with specified dimensions. In addition to the P-splines, we consider other penalty terms on the lag structure as well, as the lagged effect is assumed to fade out after some time. More specifically, varying ridge penalties can be imposed. This penalty term takes the form of either 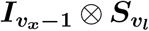 or 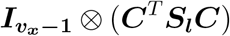 with ***S*** a diagonal matrix of dimension *v*_*l*_ and *l* respectively. More information about these varying ridge penalties can be found in Gasparrini et al. (2017).

### 3.2 An extended bivariate DLNM (2D-DLNM)

To investigate whether an interaction effect between maximum and minimum temperature is present, we propose an extension of the DLNM towards the joint effect of two variables with a delayed impact. For two delayed covariates, denoted as *x*_1_ and *x*_2_, the bivariate DLNM is defined as:

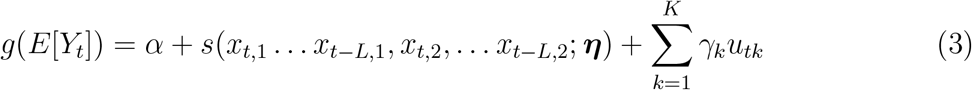

where *s* is now a smoothing function describing the joint (delayed) relationship of the variables *x*_1_ and *x*_2_, with the linear predictor. This relationship is again determined by a parameter ***η***. The other variables *u*_*k*_ are linearly related to *g*(*E*[*Y*_*t*_]) with regression coefficients *γ*_*k*_. Including the joint effect of a variable *x*_1_ and a variable *x*_2_ as well as a lag dimension, results in a three dimensional smoothing problem. Therefore, the construction of this smoothing function *s* requires an extension of the DLNM.

Analogous to the DLNM framework, we define a basis for the lag dimension to constrain the parameters along this dimension. Let 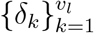 denote the basis function applied to the lag vector (0, …, *l*, … *L*)^*T*^. To allow for a flexible association between the exposures and the outcome, we choose a spline basis for the combined space of predictors *x*_1_ and *x*_2_. To this end, a tensor basis function is considered as it provides a straightforward extension of one-dimensional splines. As explained in Eilers et al. (2006), we initially require two bases for the separate exposure spaces. If 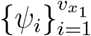 and 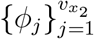 denote the univariate basis functions corresponding to *x*_1_ and *x*_2_ respectively, then 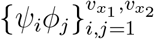 denotes the bivariate tensor spline basis. This basis can be applied to each couple of exposure variables (*x*_*t−l*,1_, *x*_*t−l*,2_).

The bivariate smoothing function can then be written as

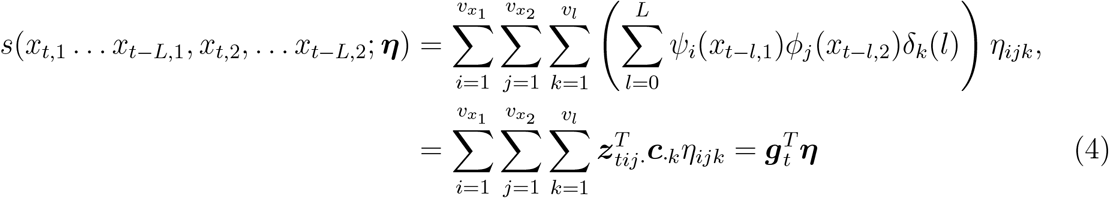

where ***z***_*tij*·_ = (*z*_*tij*0_, …, *z*_*tijL*_)^*T*^ with *z*_*tijl*_ = *ψ*_*i*_(*x*_*t−l*,1_)*ϕ*_*j*_(*x*_*t−l*,2_) and the other symbols as before.

***g***_*t*_ is now obtained by applying the 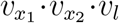 basis functions to (*x*_*t*,1_, *x*_*t*,2_). More specifically, denote by ***A***_***t***_ the 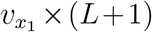 matrix obtained by *a*_*i,l*_ = *ψ*_*i*_(*x*_*t−l*,1_) and by ***B***_***t***_ the 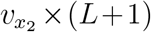 matrix containing all *b*_*j,l*_ = *ϕ*_*j*_(*x*_*t−l*,2_). Then, define the 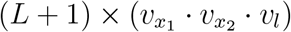 matrix:

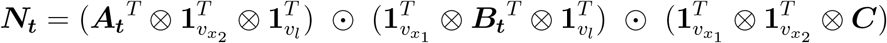

with **1** denoting vectors of ones with appropriate dimensions. The symbols ⊗ and ⊙ denote the Kronecker and Hadamard products, respectively. The vector ***g***_*t*_ is obtained by summing the matrix ***N***_***t***_ along the lag dimension (i.e. first dimension), resulting in a 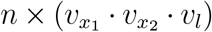 cross-basis matrix ***G*** = (***g***_1_, …, ***g***_***t***_, …, ***g***_***n***_)^*T*^.

Note that by including an intercept in 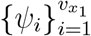 as well as 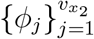, the model is again overidentified. Supposing that *ψ*_1_ and *ϕ*_1_ denote, respectively, the intercept of the basis of *x*_1_ and *x*_2_, then *z*_*t*11*l*_ = 1 for all *t* and all *l*. Hence, 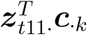 is independent of *t* for all *k* and thus perfectly correlated with the intercept. Therefore, 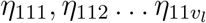 and the intercept *α* will be indeterminate. To solve this issue, we remove all terms with *i* = *j* = 1. For consistency in notation, ***η*** henceforth denotes ***η*** with coefficients belonging to terms with *i* = *j* = 1 removed. Similarly, ***G*** is obtained by removing the columns belonging to terms with *i* = *j* = 1 of matrix ***G***.

To allow the inclusion of a penalty term, P-splines are used again, leading to the following likelihood:

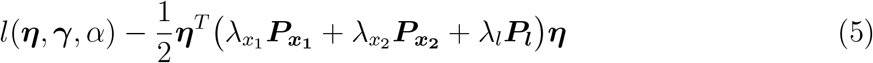

with smoothing parameters 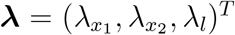. To define 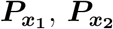 and ***P***_***l***_, we make use of the penalty matrices 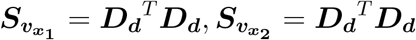 and 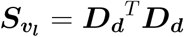, constructed from difference matrices of order *d*. The matrix 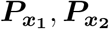 and ***P***_***l***_ can then be defined as:

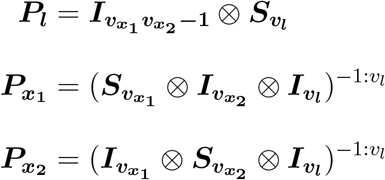

Note that the first *v*_*l*_ rows and columns are removed because of the exclusion of all terms that are collinear with the intercept. In addition to the P-splines, we can again consider varying ridge penalties in the lag dimension.

Besides from ***η, γ*** and *α*, also the smoothing parameters ***λ*** have to be estimated. To this end, different estimation methods exist within P-IRLS (penalized iterative reweighted least square). According to Wood (2011), restricted maximum likelihood (REML) is often recommended as it is less prone to severe undersmoothing as compared to alternative methods. Therefore, this method is chosen as smoothing parameter estimation method. The variance-covariance matrix of the regression coefficients is estimated using an empirical Bayes approach. The uncertainty concerning the estimation of the smoothing parameter is not taken into account in these estimates but little impact in data applications has been shown in Marra and Wood (2012).

### 3.3 Application

The previously discussed extension of the DLNM to allow the inclusion of an interaction- delayed effect is applied to our data. We fit four models to investigate the relationship between all-cause mortality counts *y*_*t*_ and maximum and/or minimum temperatures. Every model is a generalized additive model, belonging to the Poisson family. We use natural cubic splines for the year effects with 3 degrees of freedom, to control for the long term trend, natural cubic splines for the month effects with 2 degrees of freedom, to control for seasonality, indicator variables for day of the week and a linear term for winter mortality rate. A maximum lag of 14 days is used for temperature and we include population size as an offset in the model. Moreover, we include an area-specific random intercept to account for possible differences in baseline mortality risk between municipalities. Gasparrini (2022) argued that this might be important to control for structural differences between areas. Different models are fitted for women and men, as well as for people that are older than 85 and people that are between 65 and 84 years old.

Let *g* denote the gender (i.e., men or women) and *a* the age category (*>* 85 or 65 *−* 84). Denote by *i* the municipality. Then four different models (*j* = 1, …, 4) are fitted for every combination of *a* and *g*. The models are given by:

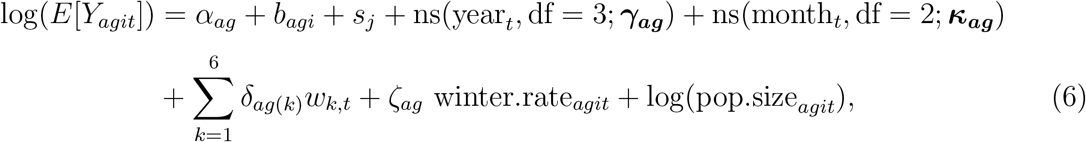

where *w*_*k*_ is an indicator of the day of the week and the random intercept 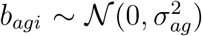 takes into account region-specific heterogeneity. Denote by *x*_1_ and *x*_2_ the maximum and minimum temperature, respectively. The smoothing function *s*_*j*_ is then constructed as:

1. *s*_1_ = *s*(*x*_*it*,1_ *x*_*i*(*t−*14),1_; ***η***_***ag*(1)**_)
2. *s*_2_ = *s*(*x*_*it*,2_ *x*_*i*(*t−*14),2_; ***η***_***ag*(2)**_)
3. *s*_3_ = *s*(*x*_*it*,1_ … *x*_*i*(*t−*14),1_; ***η***_***ag*(1)**_) + *s*(*x*_*it*,2_ *x*_*i*(*t−*14),2_; ***η***_***ag*(2)**_)
4. *s*_4_ = *s*(*x*_*it*,1_ … *x*_*i*(*t−*14),1_, *x*_*it*,2_ … *x*_*i*(*t−*14),2_; ***η***_***ag***_)

We construct the smoothing function using P-splines with 5 degrees of freedom in the lag dimension and 11 degrees of freedom for both variable dimensions (before imposing constraints). These choices are expected to provide enough flexibility in the lag as well as variable dimension. A default second order difference penalty is used in all dimensions. An additional penalty in the lag dimension is added, namely a ridge penalty ***p*** = (0, 0, 0, 1, 1)^*T*^, such that the additional *v*_*l*_ *× v*_*l*_ penalty matrix 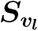 is a diagonal matrix with weights given by ***p***. It has been argued that this additional penalty is helpful in identifying an appropriate lag period, even if the specified maximum lag substantially exceeds the correct lag period (Obermeier et al., 2014). REML is used for smoothing parameter estimation.

As the Meteorological Institute of Belgium KMI reports a maximum temperature of 22.5 degrees Celsius and a minimum temperature of 13.4 degrees Celsius as ‘normal’ summer values between 1991 and 2020, these values are chosen as the temperature reference values. After model selection, we can then investigate which temperature values lead to a significant increase in relative risk (RR), as compared to the normal values. Furthermore, we calculate the attributable fraction (AF), i.e. the fraction of the variability in the outcome of interested attributed to the effect of one or more explanatory variables. Gasparrini and Leone (2014) introduce two perspectives to calculate this AF: a forward perspective, specifying the future burden expected from a given exposure, and a backward perspective, specifying the current burden resulting from a set of past exposures. Adopting a backward perspective, we examine the impact of heat during the summer periods between the year 2000 and 2019. We define a heat event as any day with both temperature measures exceeding the normal summer values i.e. the maximum temperature exceeds 22.5 degrees Celsius and the minimum temperature exceeds 13.4 degrees Celsius. The AF quantifies the proportion of deaths in people aged over 65 that can be attributed to heat during this period.

## 4. Simulation study

Before we compare the performance of these models on the data, we want to ensure that, when an interaction between maximum and minimum temperature would indeed be present, our proposed model is able to capture this trend. Therefore, we performed three simulations. In the first scenario (interaction), we simulate an interaction effect between maximum and minimum temperature, while in the second scenario (additive), there is no interaction effect. In the third scenario (benchmark), we investigate whether the models are able to identify the threshold at which relative risks start to increase when the risk remains constant across various values (i.e., there is a whole region of values with relative risk equal to 1). The predictors *x*_1_ and *x*_2_ are the maximum and minimum temperature, respectively, as observed in Flanders. For the simulation 30 randomly selected municipalities are used, with data spanning from 2000 to 2019, between May 15 and September 30. We simulated counts from a Poisson distribution, using the following model:

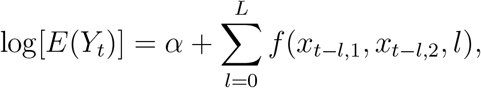

where *f* is the three-dimensional function calculating the true log relative risk, relative to a maximum and minimum temperature of 22.5 degrees Celsius and 13.4 degrees Celsius respectively.

Regarding the construction of function *f*, we considered three true underlying scenarios over lag 0 *−* 14. Scenario 1 (interaction) and Scenario 2 (additive) resemble a J-shaped temperature-mortality association with and without interactions between maximum and minimum temperature, respectively. In Scenario 3 (benchmark), the relative risk remains constant below a certain threshold value and increases for higher temperature values. An interaction between maximum and minimum temperature is again assumed. Details about these scenarios can be found in the Supplementary Materials (Web Appendix A).

Figure 1 shows a plot of the different log relative risk surfaces at lag 0. The intercept *α* is chosen such that the Pearson correlation between *E*[*Y*_*t*_] and *y*_*t*_ is approximately 0.5 in every scenario. This results in an *α* of 8.56, 7.44 and 7.50 in Scenario 1, Scenario 2 and Scenario 3 respectively.

**Figure 1:**
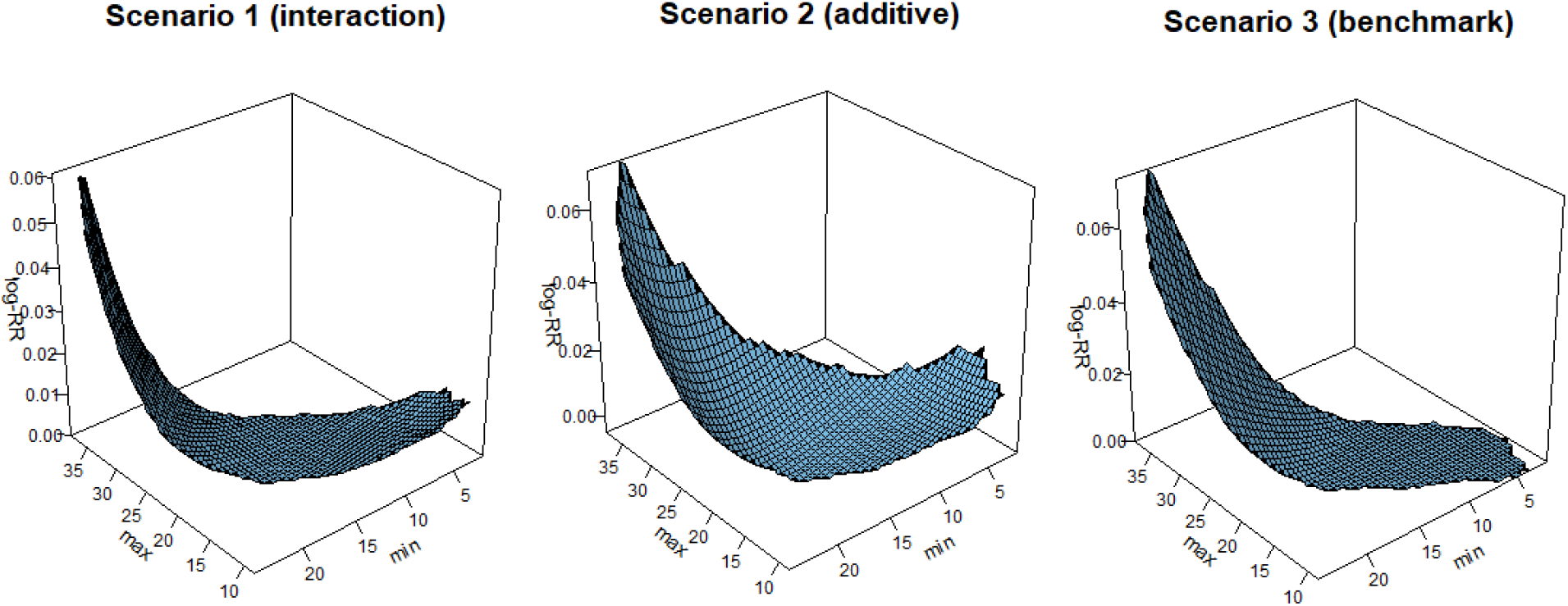
Simulated scenarios with different exposure-response associations at lag 0.

For every scenario, we compare the performance of two different estimators: namely a model with an interaction effect between maximum and minimum temperature (2D-DLNM) and a model with an additive effect of maximum and minimum temperature (DLNM). We used specifications similar to the specifications in the data applications, namely P-splines with 5 degrees of freedom in the lag dimension and 11 degrees of freedom in the variable dimension(s) (before exclusion of the collinear terms). Furthermore, the same additional ridge penalty in the lag dimension is used, namely ***p*** = (0, 0, 0, 1, 1)^*T*^. We compare the effective degrees of freedom and estimation time in every scenario. Note that the effective degrees of freedom are calculated following the approach of Wood et al. (2016), which takes into account the smoothing parameter estimation uncertainty. Moreover, we calculate the coverage of the lag-specific RR, the coverage of the overall RR (cumulative RR calculated by summing all contributions over lag 0 *−* 14) and the RMSE relative to the RMSE of the interaction estimator. We do so by calculating these values over 1000 simulations for a grid of temperature values. Taking into account the correlation between maximum temperature (ranging from 8 to 39 degrees Celsius) and minimum temperature (ranging from 1 to 26 degrees Celsius), this grid is constructed using 1114 equally spaced temperature values.

The results of the simulation study are given in Table 1. Moreover, Figure 2 shows some relevant figures. The first two rows of the figure represent the overall log RR for different maximum temperatures and a minimum temperature fixed to 20 degrees Celsius. The third and fourth row show the overall log RR for different minimum temperatures and a maximum temperature fixed to 30 degrees Celsius. Results show that in the presence of an interaction effect (Scenario 1 and Scenario 3), the additive model lacks flexibility to model this joint effect. The RMSE is higher and coverage is much lower as compared to the more flexible model that takes into account the interactions. Rows 2 and 4 of Figure 2 indicate that there are temperature regions where estimation with the additive model is much less accurate as compared to the interaction model (row 1 and row 3). Furthermore, serious undercoverage of the overall relative risk at certain minimum-maximum temperature combinations is observed with the additive model, suggesting a lack of fit (see Figure S2 in the Supplementary Materials). In the absence of an interaction effect (Scenario 2), the model allowing for interactions and the additive model perform equally well in terms of coverage, though the additive model has lower RMSE.

**Table 1:** Estimation results for the model allowing for interaction and the additive model. Given are the computation time (time in seconds), edf, lag-specific RR coverage (cov), coverage of overall RR (cov overall) and RMSE relative to the RMSE of the interaction model (RMSE).

|  |  | time | edf | cov | cov overall | RMSE |
| --- | --- | --- | --- | --- | --- | --- |
| Scenario 1 | 2D-DLNM | 110.71 | 66.63 | 0.90 | 0.96 | 1 |
|  | DLNM | 6.34 | 49.02 | 0.79 | 0.60 | 1.13 |
| Scenario 2 | 2D-DLNM | 107.21 | 51.11 | 0.93 | 0.96 | 1 |
|  | DLNM | 5.50 | 41.27 | 0.91 | 0.95 | 0.85 |
| Scenario 3 | 2D-DLNM | 112.66 | 45.06 | 0.90 | 0.94 | 1 |
|  | DLNM | 6.00 | 42.95 | 0.81 | 0.73 | 1.30 |

**Figure 2:**
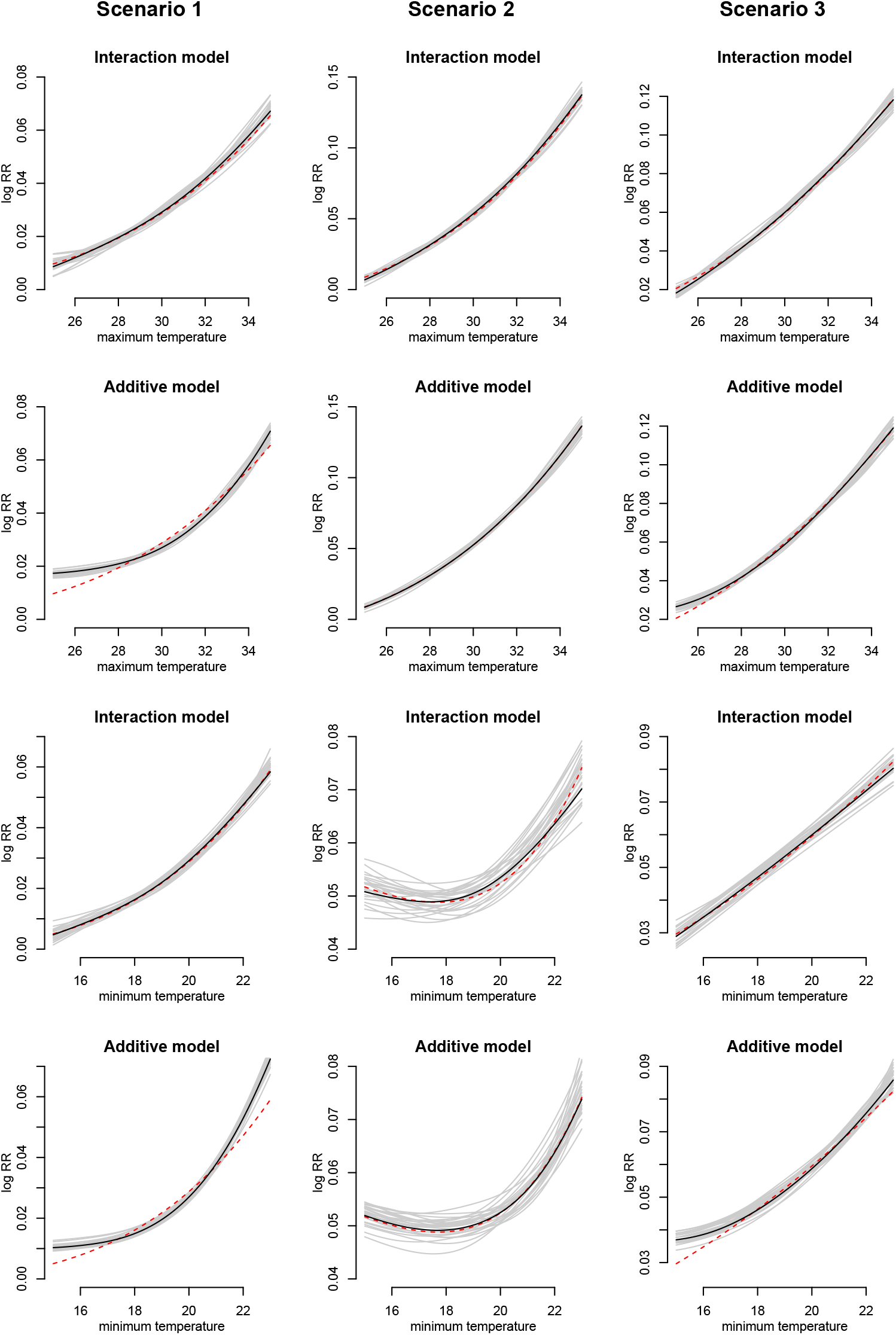
Simulation results, illustrating the performance of two estimators (Additive or interaction). The dotted red line and the solid black line represent the true log RR and the mean estimate across 1000 replicates respectively. The grey curves are the fits from 25 replicates. Rows 1-2 show the overall RR for different maximum temperatures (minimum fixed to 20) while rows 3-4 show the overall RR for different minimum temperatures (maximum fixed to 30).

In Scenario 3, interest was in identifying the threshold line beyond which there is an increased relative risk. In Figure 3, it can be seen that the threshold line can be recovered very well in the center of the grid. In this area, the relative risks of the temperatures above the true threshold line are (almost) always estimated significantly higher than 1. Close to the boundaries, the estimated relative risks of the temperatures slightly exceeding the threshold line are not significantly different from 1. However, this discrepancy is likely attributed to the wider confidence intervals. As we move further away from the centering value of 22.5 degrees Celsius maximum and 13.4 degrees Celsius minimum temperature, minor increases in risk are likely not significant due to the accompanying uncertainty. The additive model predicts a lower threshold temperature at the boundaries as compared to reality.

**Figure 3:**
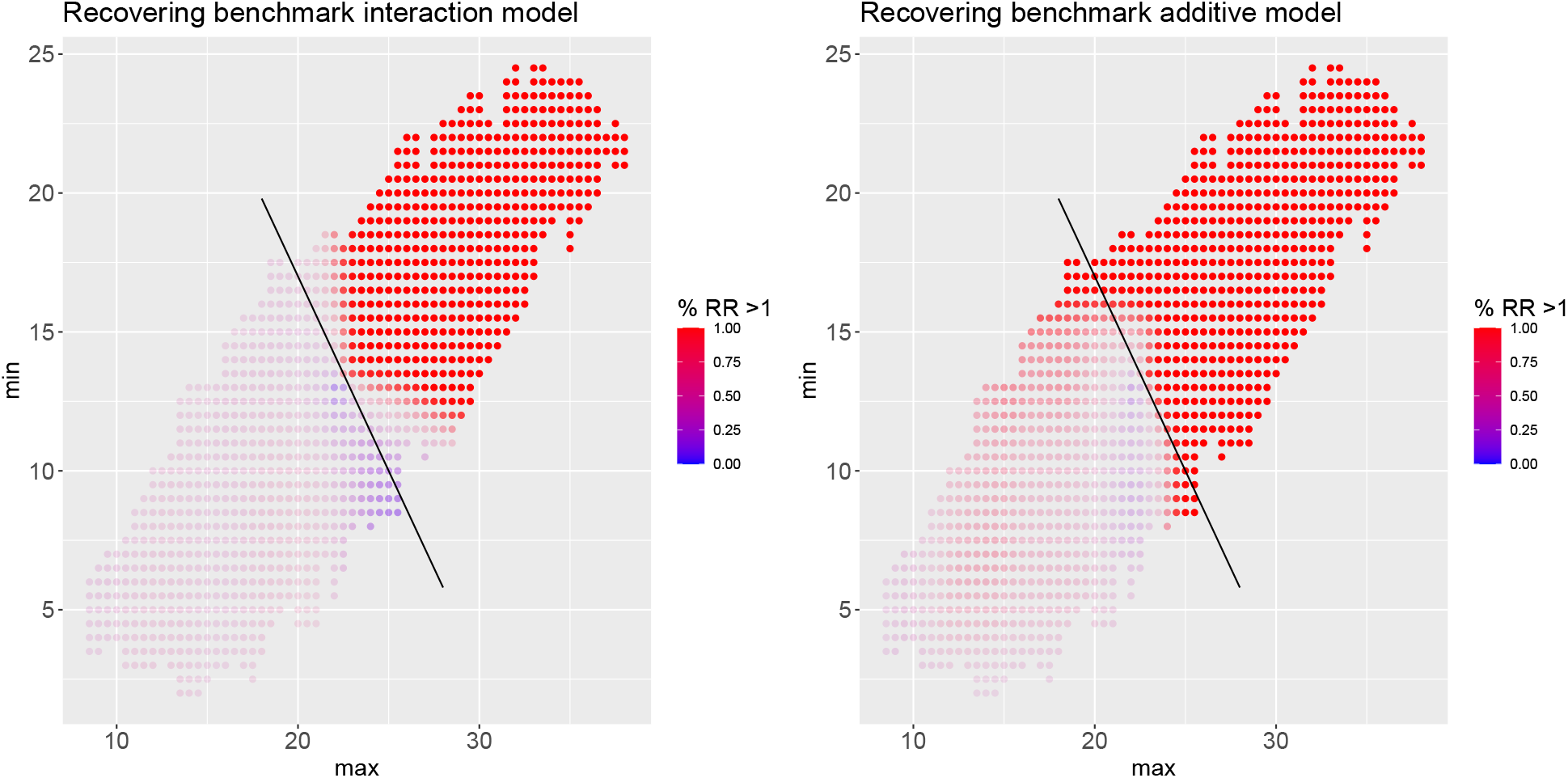
The solid black line represents the true threshold line, indicating where the relative risk exceeds 1. The colours give an indication of the percentage of the simulations for which the estimated relative risk is above 1. Opacity reflects the percentage of the simulations for which the estimated relative risk is significantly different from 1. When a color is transparent, the estimated RR is rarely, if ever, significantly different from 1. Left panel: interaction model and right panel: additive model.

## 5. Flemish mortality data results

For every age and gender group, we fitted model (6) for *j* = 1, …, 4. We compared all four models based on (conditional) AIC and the results are shown in Table 2. The effective degrees of freedom are calculated following the definition of Wood et al. (2016). For men aged over 85, the AIC comparison suggests that the fourth model, including interactions between maximum and minimum temperature, is most suitable. However, for the other strata, the additive model is preferred.

**Table 2:**
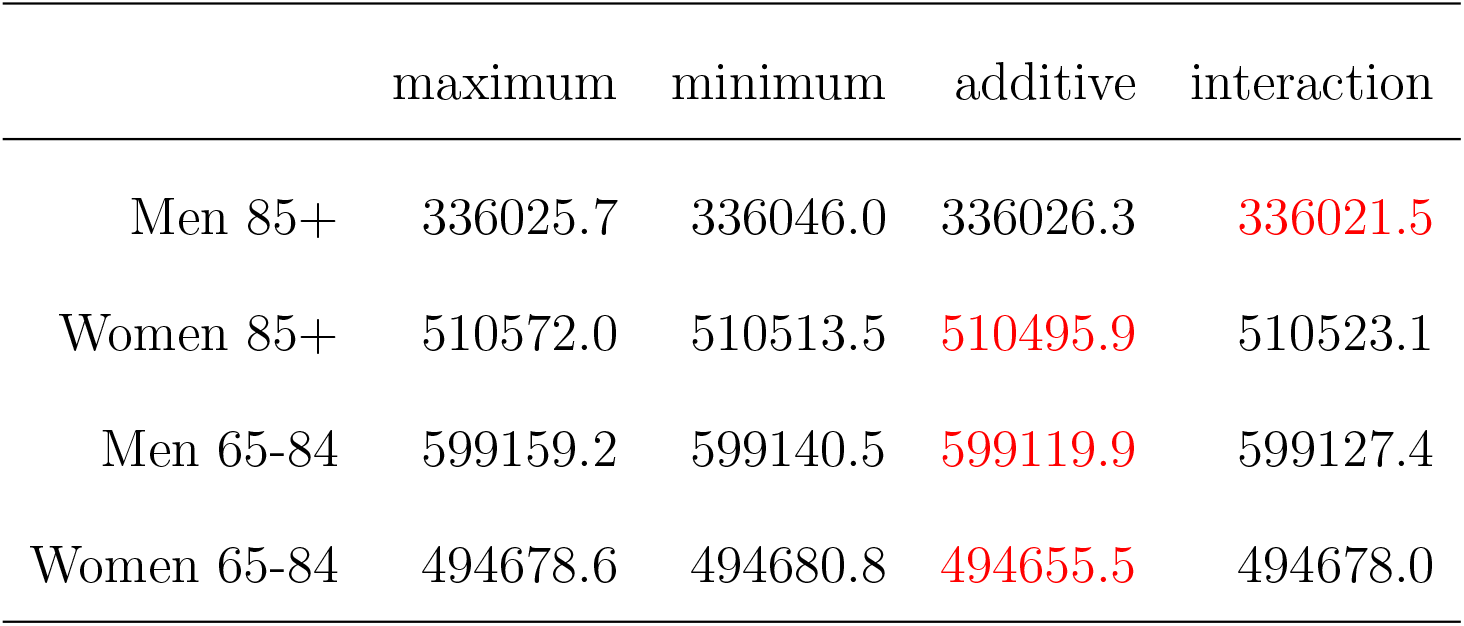
cAIC values of different models in different gender and age groups.

|  | maximum | minimum | additive | interaction |
| --- | --- | --- | --- | --- |
| Men 85+ | 336025.7 | 336046.0 | 336026.3 | **336021.5** |
| Women 85+ | 510572.0 | 510513.5 | **510495.9** | 510523.1 |
| Men 65-84 | 599159.2 | 599140.5 | **599119.9** | 599127.4 |
| Women 65-84 | 494678.6 | 494680.8 | **494655.5** | 494678.0 |

Figure 4 shows that for all age and gender groups, extreme high temperatures result in the highest estimated RR. However, differences between the groups can be observed. For men aged over 85, Figure 4 suggests that for high temperatures, relative risks are primarily influenced by maximum temperature while minimum temperature emerges as the key factor influencing risk assessment for lower temperatures. The overall risk lies significantly above 1.2 when the maximal temperature exceeds approximately 33 degrees Celsius. The relative risks for men aged between 65 and 84 years old are driven by maximum and minimum temperatures, although the minimum temperature seems to be the key factor for high temperatures. The overall risk lies significantly above 1.2 when the minimum temperature exceeds approximately 22 degrees Celsius, with the threshold only slightly depending on the maximum temperature. The relative risks for women, especially in the oldest age category, are influenced by both maximum and minimum temperature as well, as indicated by the non-vertical and non-horizontal orientation of the threshold lines. When the nighttime temperature lies above the threshold of 30 minus 35% of the day’s maximal temperature, it means it is too warm at night, with a risk of mortality significantly above 1.2, especially among the elderly. In general, we see that for women, the relative risk lies significantly above 1.2 when the maximum temperature exceeds 33 degrees Celsius or when the minimal temperature exceeds 21 degrees Celsius.

**Figure 4:**
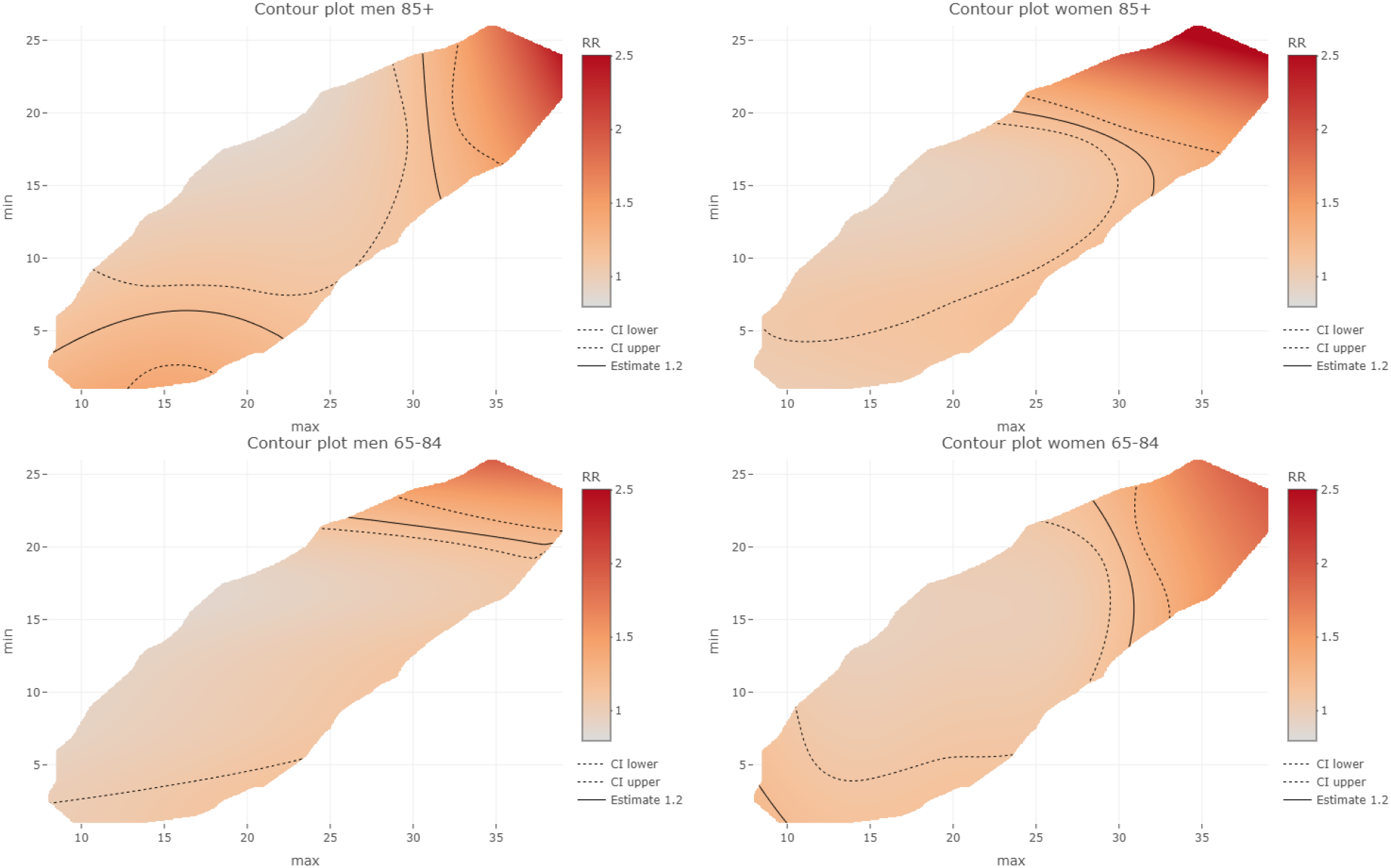
Contour plot with estimated overall RR over a lag period of 0 *−* 14 days in 4 categories of interest (UL: men 85+, UR: women 85+, LL: men 65 *−* 84 and LR: women 65 *−* 84). Maximum temperature can be found on the x-axis and minimum temperature on the y-axis. The colours give an indication of the estimated RR. The lines indicate the estimated threshold, as well as upper and lower bounds, beyond which the estimated RR exceeds 1.2.

To compare the joint effects of maximum and minimum temperatures in the four strata, we investigate the overall RR in four situations (Table 3). We look at a situation with a high maximum and minimum temperature (35 degrees Celsius and 23 degrees Celsius), a situation with high maximum but lower minimum temperature (35 degrees Celsius and 17 degrees Celsius), a situation with lower maximum and high minimum temperature (29 degrees Celsius and 23 degrees Celsius) and a situation with lower maximum and lower minimum temperature (29 degrees Celsius and 17 degrees Celsius). It is illustrated that for men above 85 and women between 65 and 84, higher maximum temperatures lead to significantly higher RR compared to lower maximum temperatures, keeping minimum temperature constant. For women aged over 85 and men aged between 65 and 84, the cooling down effect is most clearly illustrated as the overall RR is significantly lower for lower as compared to higher minimum temperatures, when the maximum temperature remains constant.

**Table 3:**
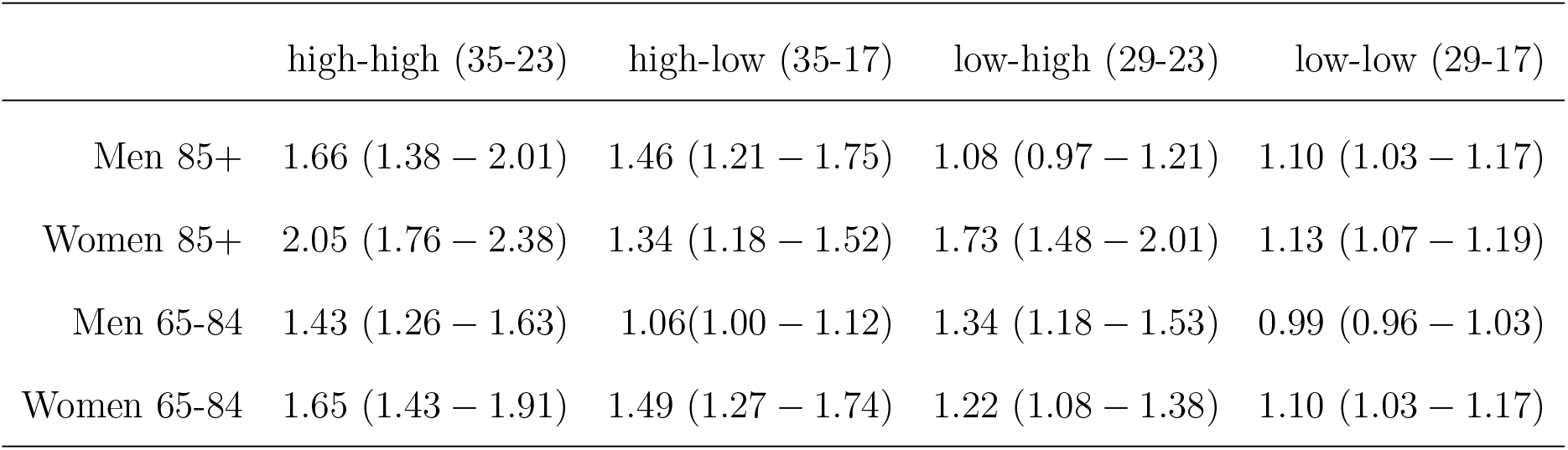
Estimated overall RR in different scenarios: (1) high maximum and high minimum temperature (35 and 23 degrees Celsius respectively), (2) high maximum and lower minimum temperature (35 and 17 degrees Celsius respectively), (3) lower maximum and high minimum temperature (29 and 23 degrees Celsius) and (4) lower maximum and lower minimum temperature (29 and 17 degrees Celsius).

|  | high-high (35-23) | high-low (35-17) | low-high (29-23) | low-low (29-17) |
| --- | --- | --- | --- | --- |
| Men 85+ | 1.66 (1.38 – 2.01) | 1.46 (1.21 – 1.75) | 1.08 (0.97 – 1.21) | 1.10 (1.03 – 1.17) |
| Women 85+ | 2.05 (1.76 – 2.38) | 1.34 (1.18 – 1.52) | 1.73 (1.48 – 2.01) | 1.13 (1.07 – 1.19) |
| Men 65-84 | 1.43 (1.26 – 1.63) | 1.06(1.00 – 1.12) | 1.34 (1.18 – 1.53) | 0.99 (0.96 – 1.03) |
| Women 65-84 | 1.65 (1.43 – 1.91) | 1.49 (1.27 – 1.74) | 1.22 (1.08 – 1.38) | 1.10 (1.03 – 1.17) |

We can compare the estimated RR in more detail for an very hot summer day with a maximum temperature of 32.38 degrees Celsius and a minimum temperature of 20.55 degrees Celsius, representing the 99th percentile of maximum and minimum temperature, respectively. The estimated overall RR for men aged over 85 is 1.32 (95%CI: 1.20 *−* 1.45) whereas for women, it is 1.51 (95%CI: 1.40 *−* 1.62). In the age category 65 until 84, we find an estimated RR of 1.16 (95%CI: 1.09 *−* 1.22) for men and 1.37 (95%CI: 1.26 *−* 1.48) for women. Hence, this temperature has a greater impact on women than men, and within each gender, its effect is more pronounced in the oldest age category. Figure 5 shows the estimated RR for the different lags in this scenario. The largest lag times are observed in the older age groups. For 85+ year olds, an effect of temperature is visible up to 7 days after a warm day. This effect is shorter among 65-84 year olds, especially males. At the 95th percentile of maximum and minimum temperature, the immediate relative risk remains highest for women aged over 85. However, the lag period is typically shorter as compared to the 99th percentile (see Figure S3 in the Supplementary Materials).

**Figure 5:**
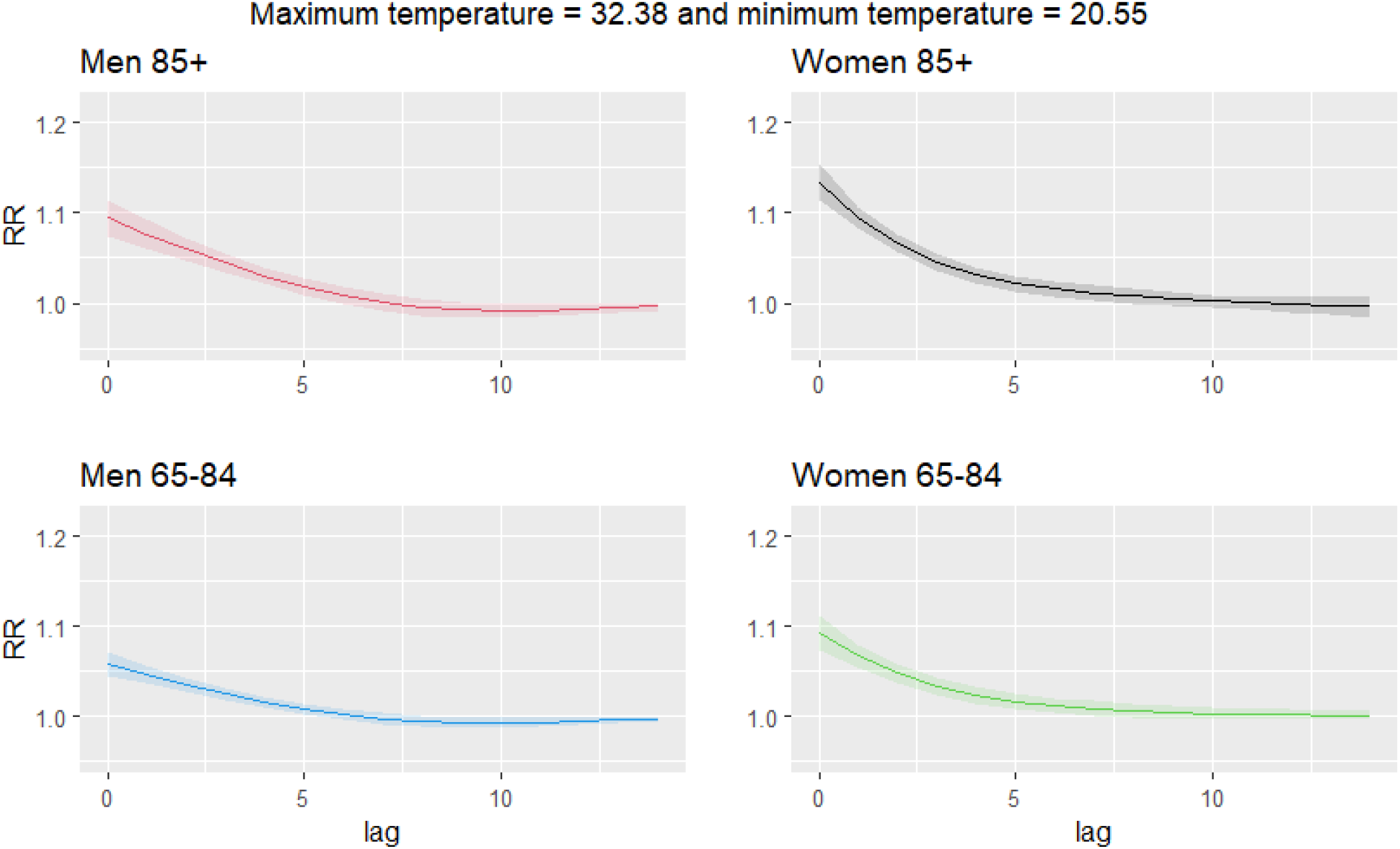
Estimated RR for a maximum temperature of 32.38 and a minimum temperature of 20.55 and different lags (UL: men 85+, UR: women 85+, LL: men 65 *−* 84 and LR: women 65 *−* 84.

The left panel of Figure 6 shows that the AF due to heat in people aged over 65, is generally lower in the eastern part of Flanders, which is the part with lower average maximum temperatures. However, this trend is not consistently gradual. Instead, variations are observed among municipalities within the same region, potentially influenced by factors beyond temperature, such as age and gender. Furthermore, it can be seen that Antwerp, having a high average minimum temperature, shows a somewhat higher estimated AF as compared to most of its surrounding areas. The right panel of Figure 6 shows a plot of the estimated AF per summer, due to heat, together with the bootstrap confidence intervals. It can be seen that the summer of 2006 has the highest AF due to heat. Although this summer ranks third in terms of average maximum temperature, it has the highest average minimum temperature. The summer of 2007, which is the summer with lowest average maximum temperature, has the lowest AF due to heat, not significantly different from zero. Although the trend in AF largely follows the temperature trend, yearly differences in the population demographics age and gender also influence the estimated AF due to heat.

**Figure 6:**
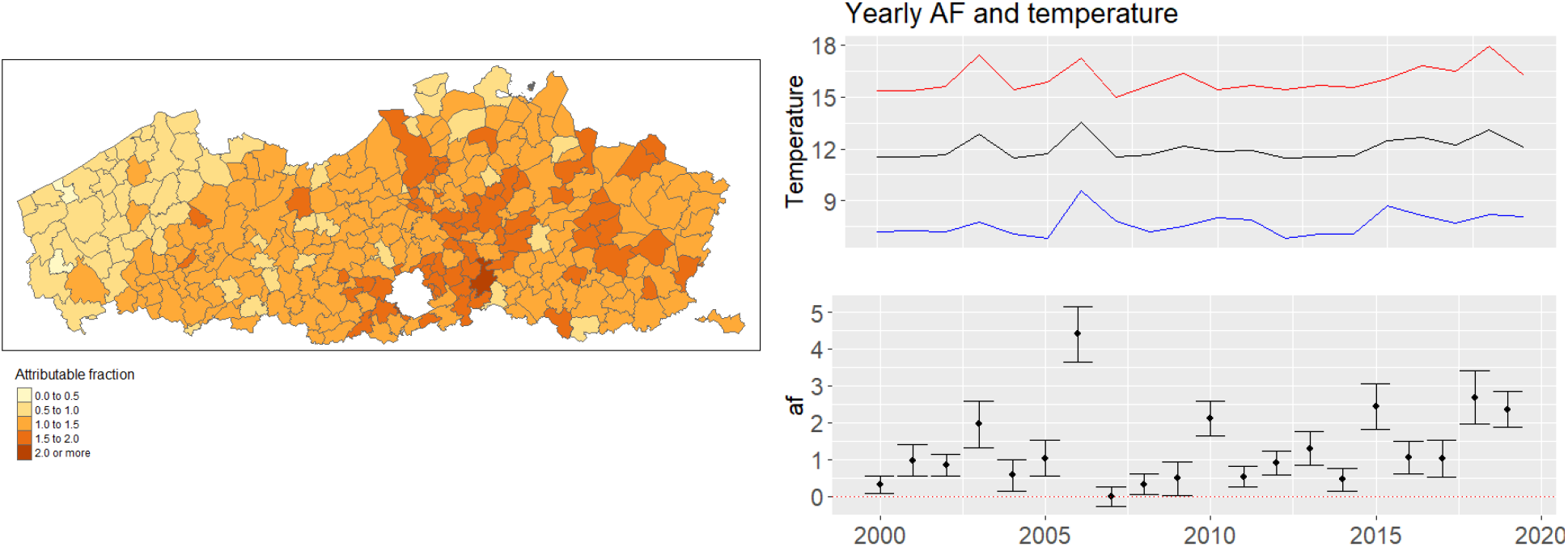
A plot of the estimated attributable fraction due to heat in the different municipalities (left panel) and the yearly total attributable fraction in Flanders due to heat with according bootstrap confidence interval, together with yearly average maximum temperature (red), minimum temperature (blue) and average temperature (black) (right panel).

In summary, the attributable fraction (AF) varies both spatially and temporally. The estimated AF due to heat over the summer periods between the year 2000 and 2019 in people aged over 65, ranges from approximately 0% to approximately 4% across different municipalities. Looking at the temporal trend, the AF shows statistically significant values in all years except for the year 2007. An estimate of the total AF due to heat in Flanders is equal to 1.31% (95%CI: 0.88% *−* 1.74%).

## 6. Discussion

In this paper, we introduced a new modelling approach that provides a flexible strategy toward the analysis of two predictors with interacting time-delayed effects. This model is an extension of the DLNM, allowing for a tri-dimensional relationship between two exposures, lag time and the response. Through the utilization of tensor splines, we construct a basis for the joint space of two predictors, subsequently combining this with the basis for the lag dimension. To address the challenge of choosing the number and placement of the knots, we make use of penalized splines.

Simulations were conducted, revealing that the conventional DLNM demonstrates limited flexibility in modeling the relationship between exposures and response, in the presence of interaction effects amongst different exposures. The flexible model introduced in this paper addresses this problem. When interaction effects are unnecessary, simulations demonstrate that the flexible model remains capable of capturing the exposure-response relationship and maintains the desirable coverage.

Using this model and comparing it with traditional parametrizations of the DLNM, we showed that both maximum and minimum temperature can have an important effect on mortality, possibly interacting. Therefore, it might be desirable to reconsider the definition of heat waves and so called ‘heat action plans’, as they are only based on maximum temperature. Based on the results of the analysis, we conclude that there is relative risk of mortality above 1.2 when the minimal temperature is above 21 degrees Celsius or the maximal temperature is above 33 degrees Celsius. While for the men above 85 years old and the women between 65 and 84, the mortality risk increases mainly if the temperature during the day is high, for men between 65 and 84 years of old, the risk is highest after warm nights. For women above 85 years old, the hotter the day, the cooler it needs to be at night to stay safe.

There are caveats to our approach, though, as it capitalises on the advanced analysis of predictors that are typically highly correlated. Nevertheless, the simulations conducted do not reveal any issues with relative risk estimation, despite a Pearson correlation of about 0.75 between the two exposure time series. Another limitation of our approach is the absence of a spatially structured random effect. We incorporated a spatially unstructured random intercept to accommodate variations in baseline risk among municipalities, yet an exploratory spatial visualisation of the results did not show a spatial pattern. Nonetheless, integrating a spatially structured random effect could potentially further improve model performance. In this paper, we calculated the relative risks compared to a chosen reference value. However, interest might be in comparing the risks to a point of overall minimum mortality. Analysis revealed that this estimated temperature often falls at the boundary of the data range. Due to the less stable estimation in this region, selecting such a value as a reference may not be suitable. Additionally, considering the uncertainty associated with estimating the point of overall minimum mortality, the computation of the confidence intervals on relative risk, would need some form of bootstrap methods. Lastly, while the present analysis is based on air temperature, data on perceived temperature could lead to valuable insights as well. This will be investigated in future research.

Despite the limitations, we believe that the flexible approach introduced in this paper presents a useful tool in analyzing potentially interacting time-delayed effects in various research fields. Although the model is designed to capture the interactions between two time-delayed exposures, extending it to more than two interacting exposures is theoretically straightforward. However, the computational feasibility of this extension remains to be explored.

## Data Availability

Data used in this study are not publicly available but can be accessed upon request from the Flemish Department of Care.

## Acknowledgements

The computational resources and services used in this work were provided by the VSC (Flemish Supercomputer Center), funded by the Research Foundation - Flanders (FWO) and the Flemish Government - department EWI.

We acknowledge the Department of Care of the Flemish government and the Flemish Institute for Technological Research -VITO for providing the data.

## References

Anderson, G. B. and Bell, M. L. (2011). Heat waves in the united states: Mortality risk during heat waves and effect modification by heat wave characteristics in 43 u.s. communities. Environmental Health Perspectives 119, 210–218.

Bao, J., Wang, Z., Yu, C., and Li, X. (2016). The influence of temperature on mortality and its lag effect: A study in four chinese cities with different latitudes. BMC Public Health 16, 375.

Besancenot, J. (2002). Vagues de chaleur et mortalité dans les grandes agglomérations urbaines. Environment Risques & Santé 1, 229–240.

Braga, A. L., Zanobetti, A., and Schwartz, J. (2001). The time course of weather-related deaths. Epidemiology (Cambridge, Mass.) 12, 662–667.

De Ridder, K., Lauwaet, D., and Maiheu, B. (2015). Urbclim-a fast urban boundary layer climate model. Urban Climate 12, 21–48.

Demoury, C., Aerts, R., Vandeninden, B., Van Schaeybroeck, B., and De Clercq, E. M. (2022). Impact of short-term exposure to extreme temperatures on mortality: A multicity study in belgium. International Journal of Environmental Research and Public Health 19, 3763.

Eilers, P. H. C., Currie, I. D., and Durbán, M. (2006). Fast and compact smoothing on large multidimensional grids. Computational Statistics and Data Analysis 50, 61–76.

Eilers, P. H. C. and Marx, B. D. (1996). Flexible smoothing with B-splines and penalties. Statistical Science 11, 89 – 121.

Gasparrini, A. (2022). A tutorial on the case time series design for small-area analysis. BMC Medical Research Methodology 22, 129.

Gasparrini, A., Armstrong, B., and Kenward, M. (2010). Distributed lag non-linear models. Statistics in Medicine 29, 2224–2234.

Gasparrini, A., Guo, Y., Hashizume, M., Lavigne, E., Zanobetti, A., Schwartz, J., Tobias, A., g, S., Rocklöv, J., Forsberg, B., Leone, M., De Sario, M., Bell, M., Guo, Y.-l., Wu, C.-f., Kan, H., Yi, S.-M., Coelho, M., Saldiva, P., and Armstrong, B. (2015). Mortality risk attributable to high and low ambient temperature: A multicountry observational study. The Lancet (London, England) 386, 369–375.

Gasparrini, A. and Leone, M. (2014). Attributable risk from distributed lag models. BMC Medical Research Methodology 14, 55.

Gasparrini, A., Scheipl, F., Armstrong, B., and Kenward, M. G. (2017). A penalized framework for distributed lag non-linear models. Biometrics 73, 938–948.

Guo, H., Du, P., Zhang, H., Zhou, Z., Zhao, M., Wang, J., Shi, X., Lin, J., Lan, Y., Xiao, X., Zheng, C., Ma, X., Liu, C., Zou, J., Yang, S., Luo, J., and Feng, X. (2022). Time series study on the effects of daily average temperature on the mortality from respiratory diseases and circulatory diseases: A case study in mianyang city. BMC Public Health 22, 1001.

Marra, G. and Wood, S. N. (2012). Coverage properties of confidence intervals for generalized additive model components. Scandinavian Journal of Statistics 39, 53–74.

Murage, P., Hajat, S., and Kovats, R. S. (2017). Effect of night-time temperatures on cause and age-specific mortality in london. Environmental epidemiology (Philadelphia, Pa.) 1, e005.

Obermeier, V., Scheipl, F., Heumann, C., Wassermann, J., and Küchenhoff, H. (2014). Flexible distributed lags for modelling earthquake data. Journal of the Royal Statistical Society Series C: Applied Statistics 64, 395–412.

Oke, T. (1982). The energetic basis of the urban heat island (symons memorial lecture, 20 may 1980). Quarterly Journal of the Royal Meteorological Society 108, 1–24.

Poelmans, L., Janssen, L., and Hambsch, L. (2022). Landgebruik en ruimtebeslag in vlaanderen, oestand 2022. VITO report (in Dutch) commissioned by Flemish Department of Environment. Tech. Rep..

Royé, D. (2017). The effects of hot nights on mortality in barcelona, spain. International journal of biometeorology 61, 2127–2140.

Scovronick, N., Sera, F., Acquaotta, F., Garzena, D., Fratianni, S., and Caradee Y. Wright, A.G. (2018). The association between ambient temperature and mortality in south africa: A time-series analysis. Environmental Research 161, 229–235.

Wei, Y., Tiwari, A. S., Li, L., Solanki, B., Sarkar, J., Mavalankar, D., and Schwartz, J. (2021). Assessing mortality risk attributable to high ambient temperatures in ahmedabad, 1987 to 2017. Environmental Research 198, 111232.

Wood, S., Pya, N., and Säfken, B. (2016). Smoothing parameter and model selection for general smooth models. Journal of the American Statistical Association 111, 1548–1563.

Wood, S. N. (2011). Fast stable restricted maximum likelihood and marginal likelihood estimation of semiparametric generalized linear models. Journal of the Royal Statistical Society. Series B (Statistical Methodology) 73, 3–36.

