## Supplementary Materials for "A penalized distributed-lag non-linear model for modeling the joint delayed effect of two predictors: impact of minimum and maximum temperature on mortality"

*Web Appendix A*

We considered the following three scenarios in the simulation study, over lag $0-14$:

- Scenario 1: a shape of the plane resembling a J-shaped temperature-mortality association with interaction between maximum and minimum temperature. In this case, the function $f(x_{t-l,1}, x_{t-l,2}, l)$ is constructed with the following functions:

  - flex1$(x_1, x_2) = \sum_{p=0}^{4} \delta_p^1 \big(0.1x_1 + 0.1x_2 + 40(0.1x_1)^{1/4}(0.1x_2)^{1/3}\big)^p$
  - flex2$(x_1, x_2) = \sum_{p=0}^{4} \delta_p^2 \big(100/(x_1 + 50) + 0.1x_2 + 0.001x_1^2 x_2\big)^p$
  - w(l) = exp(−l/2)

  such that

$$f(x_{t-l,1}, x_{t-l,2}, l) = \big((\text{flex1}(x_{t-l,1}, x_{t-l,2}) + \text{flex2}(x_{t-l,1}, x_{t-l,2})) - (\text{flex1}(22.5, 13.4) + \text{flex2}(22.5, 13.4))\big)w(l)$$

  with

$$\delta_p^1 = (-0.3813986, -0.2531853, 17.6879340, -2.7660780, 0.1127700)^T \times 10^{-6}$$

$$\delta_p^2 = (-0.95349645, -0.63296325, 0.44219835, -0.06915195, 2.81925000)^T \times 10^{-3}$$

- Scenario 2: a shape of the plane resembling a J-shaped temperature-mortality association without interaction between maximum and minimum temperature. The function $f(x_{t-l,1}, x_{t-l,2}, l)$ is constructed using:

  - fadd1$(x_1) = \sum_{p=0}^{4} \delta_p^3 x_1^p + \delta_p^4 (40 - x_1)^p$
  - fadd2$(x_2) = \sum_{p=0}^{4} \delta_p^5 (x_2 - 12)^p + \delta_p^6 (22.8 - 0.2x_2)^p$
  - w(l) = exp(−l/2)

such that

$$f(x_{t-l,1}, x_{t-l,2}, l) = \big(\text{fadd1}(x_{t-l,1}) - \text{fadd1}(22.5)\big)*w(0.75l)$$
$$+ \big(\text{fadd2}(x_{t-l,2}) - \text{fadd2}(13.4)\big)*w(l)$$

with

$$\delta_p^3 = (-2.5426572, -1.6879020, 1.1791956, -0.1844052, 0.0075180)^T \times 10^{-5}$$

$$\delta_p^4 = (-4.0682515, -2.7006432, 1.8867130, -0.2950483, 0.0120288)^T \times 10^{-6}$$

$$\delta_p^5 = (-2.118881, -0.153671, -1.406585, 0.006265, 0.982663)^T \times 10^{-6}$$

$$\delta_p^6 = (-1.6951048, -1.1252680, 0.7861304, -0.1229368, 0.0050120)^T \times 10^{-4}$$

- Scenario 3: a shape of the plane with relative risk remaining constant below a certain threshold value and increasing for higher values. An interaction between maximum and minimum temperature is assumed. The function $f(x_{t-l,1}, x_{t-l,2}, l)$ is defined by:

  – $\text{fbench}(x_1, x_2) = \begin{cases} 3.243243 \times 10^{-6}(7x_1 + 5x_2 - 225)^2, & \text{if } x_2 + 7/5x_1 \leqslant 45 \\ 0, & \text{otherwise} \end{cases}$

  – w(l) = exp(−l/2)

  and

$$f(x_{t-l,1}, x_{t-l,2}, l) = \text{fbench}(x_{t-l,1}, x_{t-l,2}) * w(l)$$

*Web Figures*

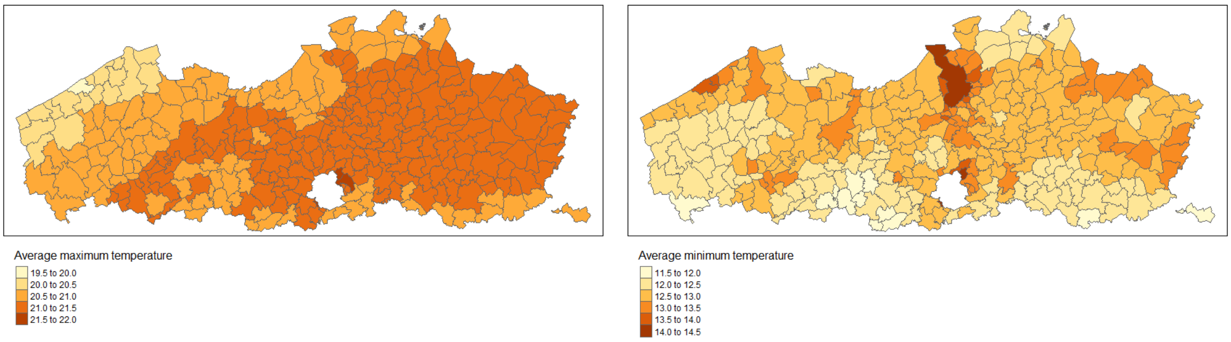


**Figure 1.** Figure S1: Average maximum (left panel) and minimum temperature (right panel) per municipality in the summer period between 2000 and 2019.

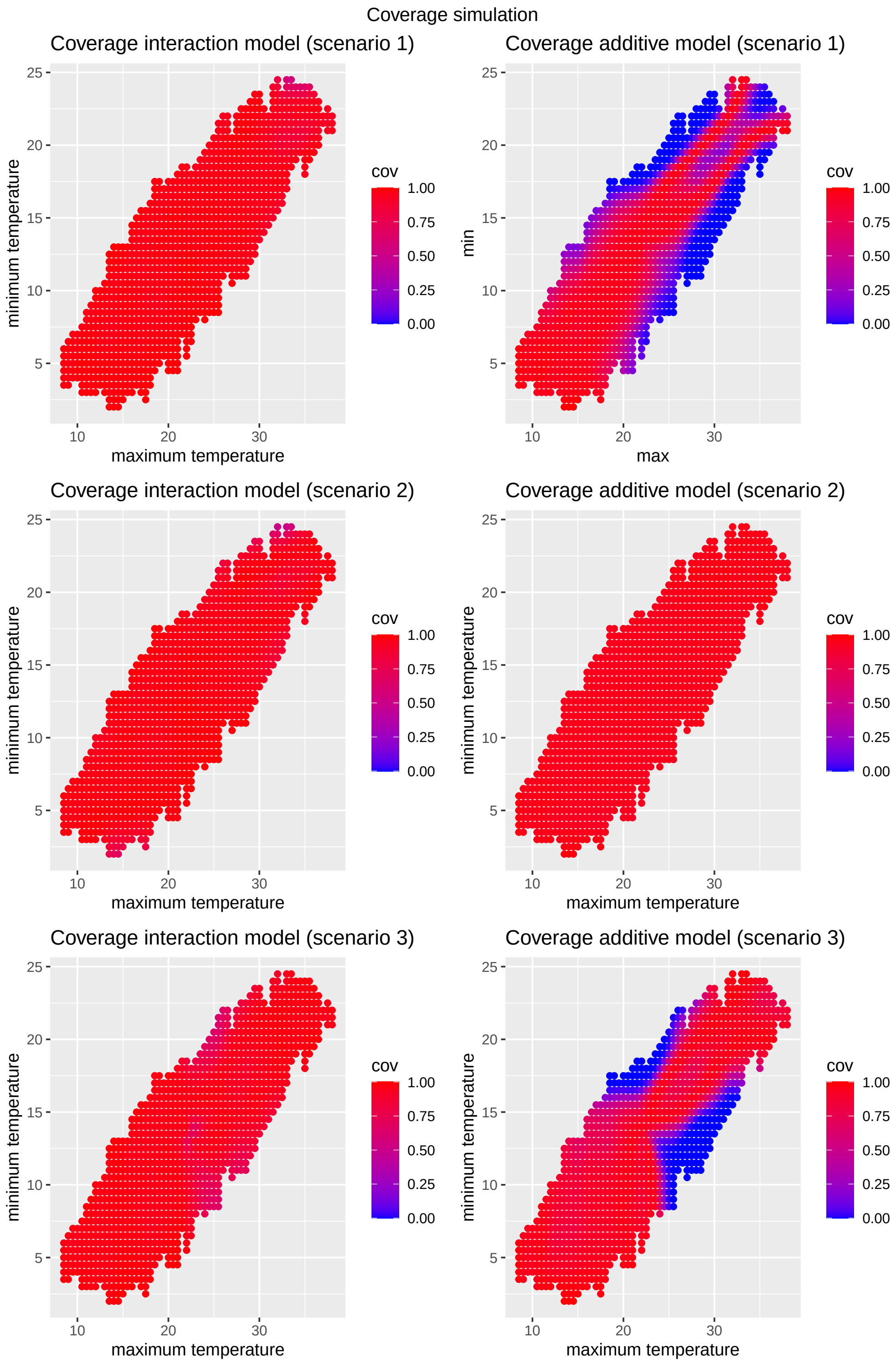


**Figure 2.** Figure S2: Coverage of overall RR for different temperature values in scenario 1 (first row), scenario 2 (second row) and scenario 3 (third row). The results using the interaction and additive model are shown in columns 1 and 2 respectively. Blue colours indicate low coverage while red colours indicate high coverage.

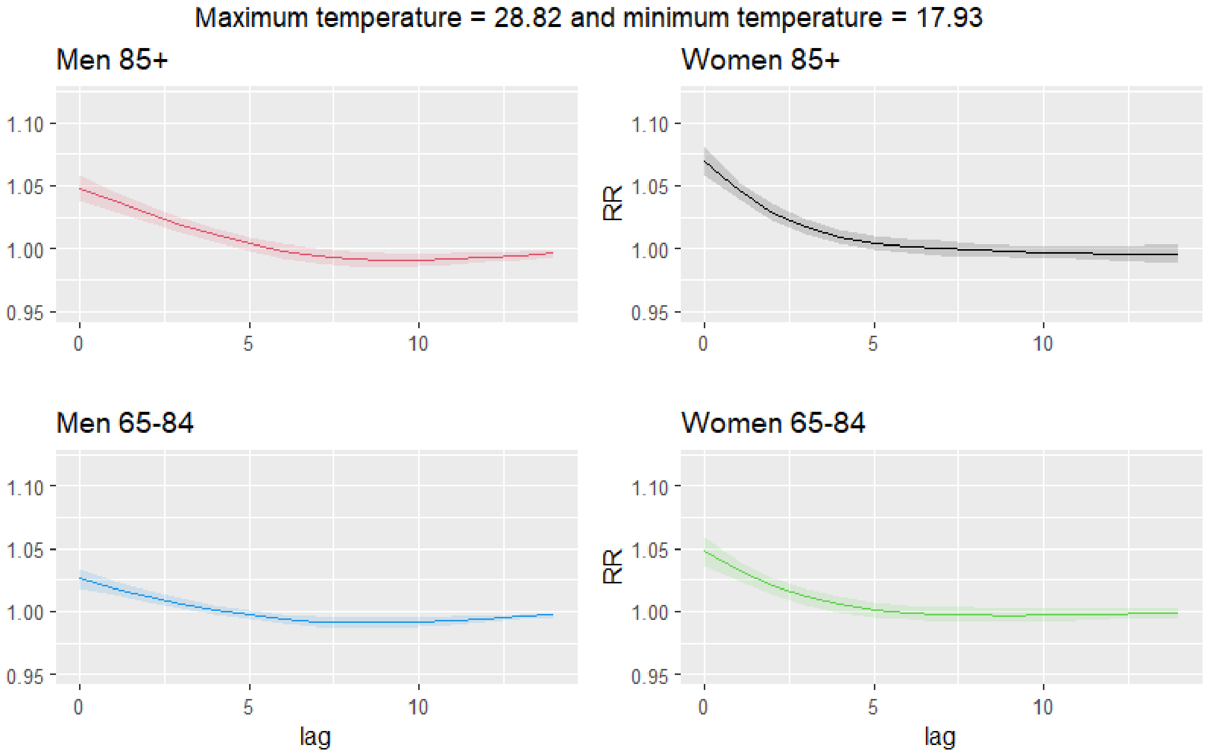


**Figure 3.** Figure S3: Estimated RR for a maximum temperature of 28.82 and a minimum temperature of 17.93 and different lags (UL: men 85+, UR: women 85+, LL: men $65-84$ and LR: women $65-84$.
